# Atrial fibrillation and kidney function: A bidirectional Mendelian randomization study

**DOI:** 10.1101/2020.07.31.20166207

**Authors:** Sehoon Park, Soojin Lee, Yaerim Kim, Yeonhee Lee, Min Woo Kang, Kwangsoo Kim, Yong Chul Kim, Seung Seok Han, Hajeong Lee, Jung Pyo Lee, Kwon Wook Joo, Chun Soo Lim, Yon Su Kim, Dong Ki Kim

**Affiliations:** Department of Biomedical Sciences, Seoul National University College of Medicine, Seoul, Korea; Department of Internal Medicine, Armed Forces Capital Hospital, Gyeonggi-do, Korea; Department of Internal Medicine, Seoul National University Hospital, Seoul, Korea; Department of Internal Medicine, Seoul National University College of Medicine, Seoul, Korea; Department of Internal Medicine, Keimyung University School of Medicine, Daegu, Korea; Biomedical Research Institute, Seoul National University Hospital, Seoul, Korea; Kidney Research Institute, Seoul National University, Seoul, Korea; Department of Internal Medicine, Seoul National University Boramae Medical Center, Seoul, Korea

**Keywords:** Atrial fibrillation, chronic kidney disease, Mendelian randomization, estimated glomerular filtration rate

## Abstract

**Aims:** To investigate the causal effects between atrial fibrillation (AF) and kidney function.

**Methods and Results:** We performed a bidirectional Mendelian randomization (MR) analysis implementing the results from large-scale genome-wide association study (GWAS) for estimated glomerular filtration rate (eGFR) by the CKDGen (N = 1,046,070) and for AF (N = 588,190) to determine genetic instruments. A bidirectional two-sample MR based on summary-level data was performed. Inverse variance weighted method was the main MR method. For replication, an allele-score based MR was performed by individual-level data within the UK Biobank cohort of white British ancestry with eGFR values (N= 321,260).

The genetical predisposition to AF was significantly associated with lower eGFR [beta - 0.002 (standard error 0.0005), P < 0.001] and higher risk of chronic kidney disease [beta 0.051 (0.012), P < 0.001], and the significance remained in various MR sensitivity analyses. The causal estimates were consistent when we limited the analysis to individuals of European ancestry. The genetically predicted eGFR did not show significant association with risk of AF [beta −0.189 (0.184), P = 0.305]. The results were similar in allele-score based MR, as allele-score for AF was significantly associated with lower eGFR [beta −0.069 (0.021), P < 0.001] but allele-score for eGFR did not show significant association with risk of AF [beta −0.001 (0.009), P = 0.907].

**Conclusions:** Our study supports that genetical predisposition to AF is a causal risk factor for kidney function impairment. However, effect from kidney function on AF was not identified in this study.

## Introduction

Atrial fibrillation (AF) is the most common heart arrhythmia disease which affects over 30 million individuals worldwide.^1^ AF is an important cause of heart failure or stroke, which leads to a higher risk of death in AF patients. Due to the global ageing trend, the prevalence of AF is predicted to further increase and the related socioeconomic burden is growing.

Chronic kidney disease (CKD) is another major comorbidity in modern medicine with increasing prevalence and large socioeconomic impact.^2^ As CKD and AF are highly prevalent in elderly individuals and share metabolic risk factors in common,^3,4^ the two comorbidities frequently coexist.^5,6^ Previous observational studies suggested that the the two diseases are risk factors for each other, suggesting a bidirectional relationship.^5,7-9^ CKD and AF are considered to synergistically aggravate the risk of stroke, adverse cardiac outcomes, and mortality.^10^

On the other hand, due to large overlap is present in risk factors and that CKD and AF commonly occurs in elderly individuals with complex comorbidities, identifying causality between CKD and AF is difficult. Thus, further evidence independent from reverse causation or confounding which is inevitable in observational studies is warranted to investigate causal effects between CKD and AF. The evidence would provide information whether modification of one may lead to a decreased risk of the other, which would help clinicians to identify targetable causal factor for AF or CKD. Furthermore, the information may guide clinicians to perform early screening for the causal factors in those with prevalent AF or CKD, preventing delayed diagnosis of important causal factors.

Mendelian randomization (MR) is a useful tool to identify causal effects from modifiable exposure on complex diseases.^11^ As MR utilizes genetic instrument, a significant association between genetic predisposition for an exposure and outcome suggests causality which is minimally affected from reverse causation or confounding. The method has been popularized in modern literature, identifying important causal factors for various complex diseases.^12-14^

In this study, we performed a bidirectional MR analysis between AF and kidney function by utilizing the recent large-scale meta-analysis genome-wide association studies (GWAS). We hypothesized that a directional causal effect of AF or kidney function may be present for each other. We both performed a two-sample MR based on summary-level data and an allele-score based MR with individual level-data. Throughout the analyses, we identified a consistent result suggesting causal effect of atrial fibrillation on kidney function impairment.

## Methods

### Ethical considerations

The study was performed in accordance with the Declaration of Helsinki. The study was approved by the institutional review boards of Seoul National University Hospital (No. E-1910-044-1067) and the UK Biobank consortium (application No. 53799). The genetic instrument implemented in this study has been previously published,^15,16^ and summary-level data were utilized. The summary statistics for the CKDGen genome-wide association study (GWAS) meta-analysis for kidney function traits are in the public domain (URL: https://ckdgen.imbi.uni-freiburg.de/). The summary statistics for AF trait are available on the Cardiovascular Disease Knowledge Portal (URL: https://www.braodcvdi.org/). As the study investigated anonymous database or summary-level data, requirement for informed consent was waived.

### GWAS meta-analysis for AF

We implemented the previous multi-ethnic GWAS meta-analysis for AF for this study.^16^ The study investigated more than 588,190 individuals (65,446 AF cases) mainly from AFGen, Broad AF, UK Biobank, and Biobank Japan study, including 84.2% of individuals from European ancestry, 12.5% from East Asian, 2% from African-Americans, and 1.3% from Brazilian and Hispanics. The study reported 94 independent AF-associated loci reaching genome-wide significance with AF trait by combined-ancestry meta-analysis. Additional GWAS for individuals of European ancestry reported 84 independent SNPs that were in genome-wide significant association with AF. The identified genetic variants implicate genes enriched within AF-related pathways, including cardiac development, electrophysiologic mechanism, and contractile or structural functions. We utilized the reported SNPs as the genetic instrument for AF in this study. In addition, the transethnic GWAS meta-analysis summary statistics were available through the Cardiovascular Disease Knowledge Portal, and we used the data as the outcome summary statistics for AF in our two-sample MR analysis.

### GWAS meta-analysis for kidney function

CKDGen provides the largest GWAS meta-analysis results for kidney function traits to date.^15^ The study meta-analyzed 121 GWAS including 765,348 individuals, with 74.1% of European ancestry, 23.4% East or South Asians, 1.8% of African-American, and 0.6% of Hispanics. The prevalence of CKD stage 3 or higher was approximately 8%. The study performed additional replication with Million Veterans Program data and reported 264 SNPs that reached genome-wide significance level for estimated glomerular filtration rate (eGFR) determined by serum creatinine values, a widely used marker for kidney function, from the multi-ethnic GWAS meta-analysis. In addition, the study performed GWAS limited to European ancestry individuals, reporting 256 independent SNPs. The genetic variants reported in the study implicated genes expressed in kidney related tissues. We utilized the SNPs as the genetic instrument for this study and downloaded the summary statistics, both for trans-ethnic results and results within the European ancestry individuals, from the CKDGen and implemented as the outcome summary statistics for our two-sample MR analysis.

### UK Biobank data

We utilized the UK Biobank data to perform additional GWAS to identify confounder associated SNPs and trim the genetic instrument. Also, the allele-score based MR was performed with the data. The UK Biobank is one of the largest prospective cohort including > 500,000 individuals with age 40-69 across the United Kingdom, which the details have been published previously.^17-19^ The data is extensively phenotyped and genotyped, thus, identifying confounder information in the individual-level data was possible. Moreover, the study measured baseline serum cystatin C, which is a superior biomarker for kidney function than serum creatinine,^20^ thus, we trimmed the genetic instrument for eGFR with the information. The details of the collected data from the UK Biobank are described in the Supplemental Methods.

### Methods to determine genetic instrument

First, we performed the analysis with the total reported SNPs from the implemented studies throughout the study. Second, we trimmed the genetic instrument to avoid bias from confounding, as the causal estimates should not be biased by a genetic variant that is strongly correlated to other possible confounders to reveal causal effects in MR analysis. We performed GWAS with the UK Biobank data adjusted for age, sex, age×sex, age^2^, and the first 10 principal components of the genetic information, for phenotypes of obesity, hypertension, dyslipidemia, and diabetes mellitus, the components of metabolic syndrome, in the UK Biobank data. The GWAS was performed with 406,733 individuals those were not with excess kinship, outliers by heterozygosity or missing rates, without sex chromosome aneuploidy. When performing analysis with results from European ancestry population, the GWAS was limited to 337,138 individuals of white British ancestry in the UK Biobank passing the quality control. We introduced a threshold to robustly exclude confounder associated SNPs and the SNPs not reaching P < 1×10^−5^ level of association with the confounders remained in the trimmed genetic instrument. For the genetic instrument for eGFR, as serum creatinine levels can be biased from diet or body shape, we additionally excluded SNPs that not reached Bonferroni-adjusted significance level (0.05/tested number of SNPs) association with the eGFR values determined by the CKD-EPI equation based on cystatin C levels.^21^ In addition, we implemented another previously reported genetic instrument which showed significant causal effects from kidney function to blood pressure, also from the CKDGen study, to additionally test the causal effects of kidney function on AF.^22^ The genetic instrument was similarly included SNPs that were identified from the CKDGen study within European ancestry individuals but was more stringently trimmed based on genetic information and literature review by supervised manner.

### Two-sample MR based on summary-level data

The two-sample bidirectional MR was first performed with trans-ethnic summary level data and the causal estimates between AF and CKD were identified (Figure 1). In concerns of overlapping samples in East Asian population (e.g. Biobank Japan), additional analysis performed within the data from European ancestry individuals, testing the causal estimates from AF to kidney function traits. However, the causal estimates from eGFR to AF within the European ancestry samples were not performed by this summary-level MR as the summary statistics, standard error, for individuals of European ancestry was unavailable. In summary-level MR, the SNPs not overlapping between the summary statistics or that were palindromic with intermediate allele frequencies were disregarded.^23^ The main method for the two-sample MR was the fixed-effects inverse variance weighted method. Sensitivity analyses were performed with widely recognized MR sensitivity analysis methods. First, MR-Egger regression, which yields pleiotropy-robust causal estimates, was performed, with bootstrapped standard errors.^24^ Second, the panelized weighted median mode method was implemented which derives valid causal estimates even in conditions when invalid instruments are present.^25^ Finally, MR-PRESSO was performed, which detect and correct the effects from outliers, yielding causal estimates that are robust to heterogeneity.^26^ The two-sample MR analysis was performed by the TwoSampleMR package in R (version 4.0.2, the R foundation).^27^

**Figure 1.**
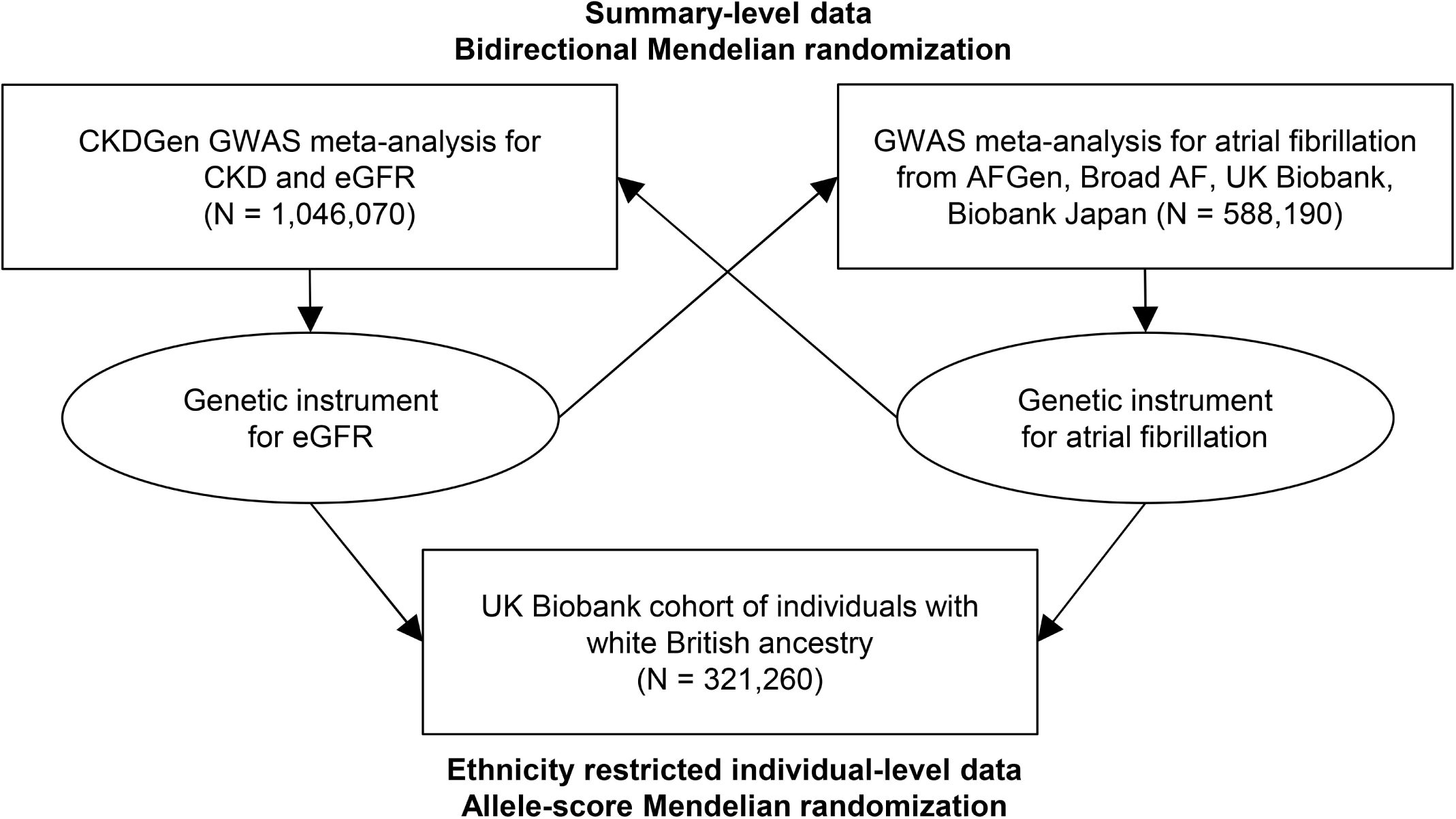
Study flow diagram. The study consisted of two parts; a MR analysis based on summary-level data and an allele-score MR analysis based on individual-level data. eGFR = estimated glomerular filtration rate.

### Allele-score MR based on individual-level data

To replicate the findings, allele-score MR based of individual-level data was performed and restricted the analysis to data from European ancestry individuals to decrease the effects from multi-ethnicity (Figure 1).^28^ Within the UK Biobank data, 321,260 individuals of white British ancestry passing quality control for genetic information and with baseline eGFR values were included for the individual-level MR. The kidney function outcome was eGFR determined by CKD-EPI equation based on baseline serum creatinine values, and CKD outcome was not tested as the UK Biobank cohort has very low prevalence of CKD due to healthy volunteer bias.^17^ The AF outcome was included from the hospital admission records or main causes of death, identified by ICD-10 diagnostic codes of I48 or ICD-9 diagnostic codes of 4273. We calculated allele scores for the exposures by multiplying the gene dosage matrix with the effect sizes of the genetic instrument, in GWAS summary statistics within European ancestry individuals, by using PLINK 2.0 (version alpha 2.3).^29^ The association between genetically predicted eGFR or AF with phenotypical AF or eGFR was tested by linear or logistic regression analysis, adjusted for age, sex, genotype measurement batch, and the first 10 principal components.^13^

## Results

### MR results from summary-level data

Of the 94 SNPs for AF in the trans-ancestry GWAS meta-analysis, one SNP did not overlap with the summary statistics in the CKDGen study, and one SNP was removed for being palindromic with intermediate allele frequencies. The genetically predicted AF by 92 SNPs (Supplemental Table 1) was significantly associated higher risk of CKD and lower eGFR (Figure 2 and Table 1). The causal estimates remained significant with all implemented MR methods. After reducing the number of SNPs to 86 in the genetic instrument by excluding confounder-associated variants (Supplemental Table 2), genetical predisposition for AF remained to be significantly associated with higher risk of CKD or lower eGFR. Furthermore, when we limited the analysis from the data from individuals of European ancestry (Supplemental Table 3), the genetically predicted AF, by 73 SNPs (Supplemental Table 4), was still significantly associated with higher risk of CKD with all implemented MR methods. The causal estimates for eGFR was marginal, but after removing the confounder-associated SNPs (Supplemental Table 5), the genetical predisposition to AF was still significantly associated with higher risk of CKD and lower eGFR.

**Figure 2.**
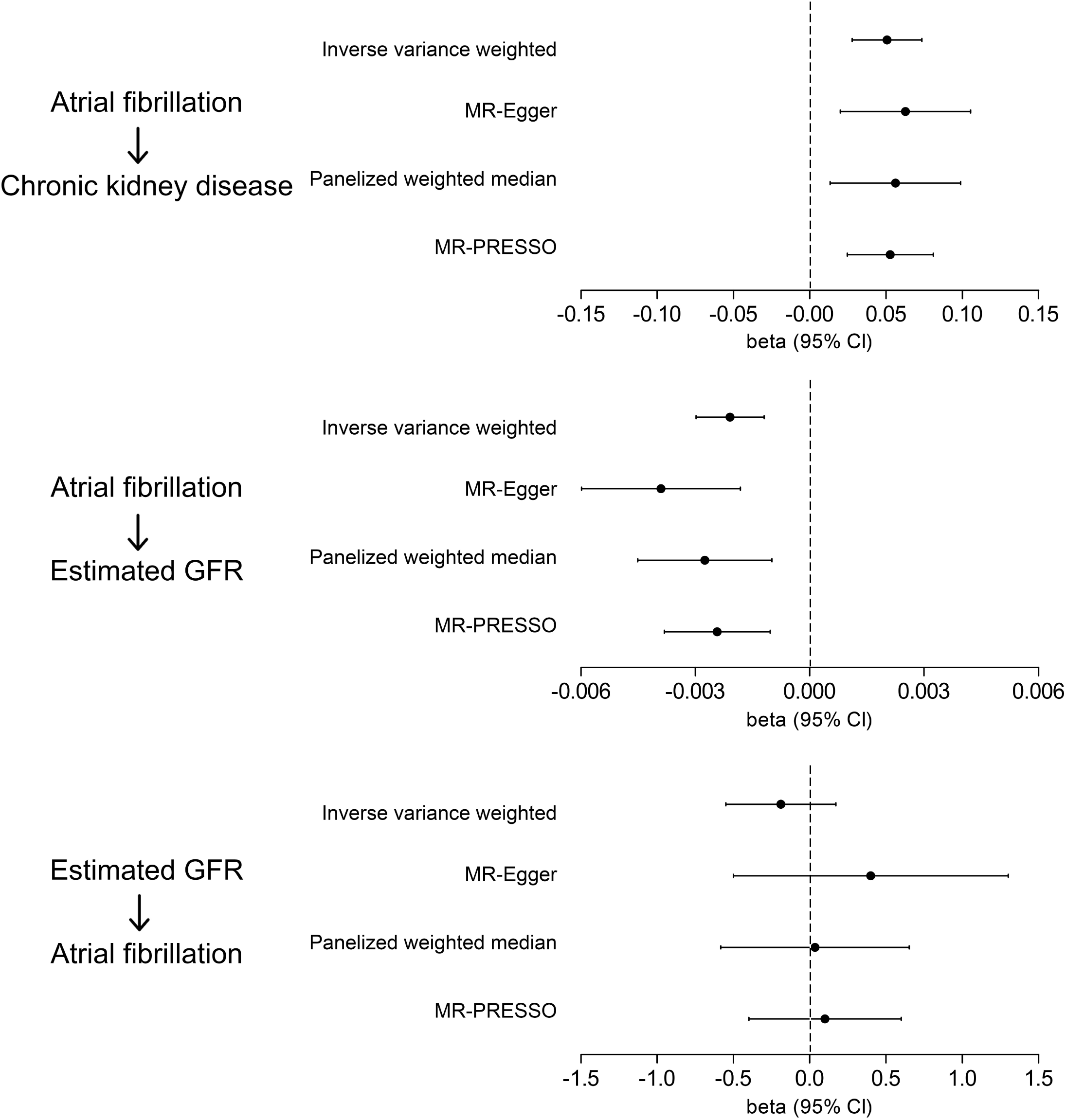
Two-sample MR analysis resulted showing causal estimates from genetic predisposition for atrial fibrillation or estimated glomerular filtration on the outcomes. The arrows indicate the tested direction of the causal estimates. The x-axes indicate beta coefficients from the two-sample MR analysis and 95% confidence intervals (± 1.96 × standard error). The results are from total SNPs of the genetic instrument. GFR = glomerular filtration rate.

**Table 1.** Causal estimates from the summary-level data-based MR from the trans-ethnic GWAS summary statistics.

| Genetically predicted exposure | Outcome | MR method | With total SNPs |  |  | With trimmed genetic instrument |  |  |
| --- | --- | --- | --- | --- | --- | --- | --- | --- |
|  |  |  | beta | standard error | P | beta | standard error | P |
| Atrial fibrillation | eGFR | Inverse variance weighted | -0.002 | 0.0005 | < 0.001 | -0.002 | 0.0005 | < 0.001 |
|  |  | MR-Egger | -0.004 | 0.001 | < 0.001 | -0.004 | 0.0010 | < 0.001 |
|  |  | Panelized weighted median | -0.003 | 0.0009 | 0.002 | -0.003 | 0.0009 | 0.002 |
|  |  | MR-PRESSO | -0.002 | 0.0007 | 0.001 | -0.002 | 0.0007 | 0.001 |
| Atrial fibrillation | CKD | Inverse variance weighted | 0.051 | 0.012 | < 0.001 | 0.053 | 0.0119 | < 0.001 |
|  |  | MR-Egger | 0.063 | 0.022 | < 0.001 | 0.066 | 0.0215 | < 0.001 |
|  |  | Panelized weighted median | 0.056 | 0.022 | 0.010 | 0.056 | 0.0216 | 0.002 |
|  |  | MR-PRESSO | 0.053 | 0.014 | < 0.001 | NA<sup>a</sup> | NA<sup>a</sup> | NA<sup>a</sup> |
| eGFR | Atrial fibrillation | Inverse variance weighted | -0.189 | 0.184 | 0.305 | 0.033 | 0.245 | 0.892 |
|  |  | MR-Egger | 0.401 | 0.460 | 0.189 | 1.763 | 0.606 | 0.002 |
|  |  | Panelized weighted median | 0.035 | 0.315 | 0.911 | 0.563 | 0.394 | 0.153 |
|  |  | MR-PRESSO | 0.101 | 0.255 | 0.692 | 0.468 | 0.314 | 0.139 |

MR = Mendelian randomization, SNP = single nucleotide polymorphism, eGFR = estimated glomerular filtration rate, CKD = chronic kidney disease
<sup>a</sup> MR-PRESSO was not performed here as MR-PRESSO global test did not identify significant heterogeneity among the genetic instrument and no correction was necessary.

On the other hand, genetically predicted eGFR, by 212 SNPs (Supplemental Table 6), as 48 SNPs were palindromic and 4 SNPs did not overlap, did not show consistently significant association with risk of CKD and the directions of the causal estimates were also inconsistent. After trimming the genetic instrument (Supplemental Table 7), the results by 90 SNPs remained similar, and the causal estimates showed inconsistent results.

### Descriptive statistics of the individual-level dataset

Among the 321260 individuals of white British ancestry, median age was 58 (interquartile ranges: 51-63) years old, including 46.3% males and 53.7% females. The number of individuals with AF was 14463 (4.5%) and their median eGFR was 92.5 (82.6-99.5) mL/min/1.73 m^2^. The prevalence of CKD stage 3-5 was 2.3%. The explained variance of eGFR by the allele scores for eGFR was 3.7% (multiple R-squared method) by the total SNPs and was 2.1% by the trimmed genetic instrument within the individuals of European ancestry. The explained variance of AF by the allele scores for AF was 2.4% (McFadden’s pseudo-R square) by the total SNPs and was again 2.4% by the trimmed genetic instrument.

### MR results from individual-level data

The allele-scores for AF based on the genetic instrument developed from the European ancestry population was significantly associated with lower eGFR in the UK Biobank (Table 2). The association remained significant even after excluding SNPs that were strongly associated with possible confounders.

**Table 2.** Causal estimates from the individual-level data based MR in the white British individuals of the UK Biobank.

| Genetically predicted exposure | Outcome | With total SNPs |  |  | With trimmed genetic instrument |  |  |
| --- | --- | --- | --- | --- | --- | --- | --- |
|  |  | beta | standard error | P | beta | standard error | P |
| Atrial fibrillation | eGFR | -0.069 | 0.021 | < 0.001 | -0.079 | 0.021 | < 0.001 |
| eGFR | Atrial fibrillation | -0.001 | 0.009 | 0.907 | -0.0001 | 0.009 | 0.986 |

SNP = single nucleotide polymorphism, eGFR = estimated glomerular filtration rate
Effect sizes of a standard deviation increase in the allele-scores were shown. The causal estimates were adjusted for age, sex, genotype measurement batch, and the first 10 principal components of the genetic information in the UK Biobank data.

However, the allele scores for eGFR did not show significant association with AF outcome. The association was insignificant even after we recalculated the allele-scores by trimming the genetic instrument by disregarding 139 SNPs from the genetic instrument for eGFR (Supplemental Table 8). Further, when we introduced the genetic instrument which was robustly filtered in a previous study, the allele scores, based on 33 SNPs, still did not show significant association with the risk of AF (Supplemental Table 9).

## Discussion

In this MR analysis, significant causal effects from AF to kidney function impairment has been demonstrated by utilizing the largest GWAS results to date for AF and kidney function. On the other hand, genetically predicted kidney function was insignificantly associated with AF, although the genetic instrument for eGFR explained similar variance as the genetic instrument for AF. Our study suggests that AF causally aggravates kidney function, but a direct effect from kidney function on AF remains uncertain.

Close association between CKD and AF has been reported repetitively by observational studies.^5-8,10^ As the burden of CKD and AF would increase in the future along with the ageing issue, treatment of AF in CKD patients has been an important issue because appropriate anticoagulation may be different in CKD population when compared to healthy individuals.^30^ Moreover, other studies focused on that the bidirectional relationship between CKD and AF, and the diseases were suspected to increase the risk of each other.^5-7,9,10^ However, assessment of direct causality between CKD and AF was difficult, as undiagnosed AF or CKD is possible,^31,32^ the disorders share many risk factors raising the concerns of confounding, and issue of reverse causality cannot be disregarded in observational studies.

By this study, we assessed the bidirectional causal effect between CKD and AF by implementing MR analysis. As MR tests the effect from genetical predisposition to an exposure, which precedes any confounders or outcome occurrence, MR can demonstrate causal effects but requires to meet three assumptions.^11^ First, the relevance assumption, which means the genetic instrument should be strongly associated with the exposure of interest, which was fulfilled by the previous GWAS meta-analysis which robustly provided the genetic variants with strong association with the exposures. Second, the restriction-exclusion assumption, which means the effect should be through the exposure of interest. Although a direct test for this assumption is impossible, it would be plausible to assess the causal effects between the two closely associated comorbidities. Third, the independence assumption, which means that the genetic variant in the instrument should not affect the outcome through a confounder (horizontal pleiotropy). We performed multiple pleiotropy-robust MR methods and disregarded variants that might have horizontal pleiotropic effect on our results.^24-26^ Thus, by performing extensive MR analysis, our study suggested that AF causally increases the risk of kidney function impairment, on the other hand, the causal effects from kidney function on AF were insignificant.

Clinical application of our study can be suggested as below. First, as AF is a causal risk factor for CKD, appropriate management of AF may lead to reduce risks of kidney function impairment. Thus, a future clinical trial may assess kidney function outcomes when performing interventions for AF. Second, as AF is one of the causal factors for CKD, clinicians may perform early screening of AF in CKD patients.^31^ Particularly, those with common risk factors for AF or with old age may be the target group that warrants such strategy, and such early diagnosis may lead to a reduced risk of progression of kidney function impairment. Third, clinicians may monitor kidney function in AF patients as they burden a higher risk of CKD due to their presence AF.

That kidney function did not show significant causal effects for AF should be interpreted in caution. Our study suggests that effects from AF to CKD may be prioritized than the effects of reverse direction in general people. However, as genetic instrument explains a portion of a phenotype, a possibility remains that a modest degree of effect may be present from kidney function on AF. Also, as we tested the effects mostly from mild-to-moderate CKD, the cardiovascular effects from severe kidney failure or acute kidney injury may be different from the current findings. Moreover, as CKD and AF share risk factors, certain CKD events would still precede AF.^8^ However, considering that kidney function also did not show significant causal effects to heart failure in a previous study implementing MR,^33^ the direct effect from kidney function on AF and heart failure would be smaller than those from other important causal factors for AF.

There are several limitations in this study. First, the two-sample MR utilizing the trans-ethnic summary statistics (e.g. Biobank Japan) and allele-score based MR (e.g. UK Biobank) have overlap in samples in genetic instrument development and outcome assessment. Although the findings remained significant with independent individuals of European ancestry, that overlap in samples in two-sample MR may cause bias should be noted.^34^ Second, although the causal effects from AF to CKD was demonstrated, whether the risk of kidney function impairment can be actually reduced through management of AF should be proven in a future trial. Third, as genetic analysis is weak to detect non-linear effects, the effects from profound kidney function impairment or mild AF may be different from our findings.

In conclusion, AF is a causal risk factor for kidney function impairment. Clinicians may pay attention to AF in CKD patients and monitor kidney function in those with AF. A further study may consider whether appropriate management of AF may lead to decreased risk kidney function impairment.

## Data Availability

The data underlying this article were accessed from the CKDGen consortium (URL: https://ckdgen.imbi.uni-freiburg.de/) the Cardiovascular Disease Knowledge Portal (URL: https://www.braodcvdi.org/). The UK Biobank data for this study will be made available in the UK Biobank consortium (biobank.ctsu.ox.ac.uk).

## Acknowledgements

The study was based on the data provided by the UK Biobank consortium (application No. 53799). We thank the investigators of the previous studies who provided valuable genetic summary statistics for this study.

## Supplemental Material Table of Contents

### Supplemental Methods

**Supplemental Table 1. Summary statistics of 92 SNPs that were included in the genetic instrument for atrial fibrillation in the summary-level MR**.

**Supplemental Table 2. Association between SNPs included in the genetic instrument for atrial fibrillation and the possible confounders in the UK Biobank data**.

**Supplemental Table 3. Summary-level data-based MR for the estimates of causal effects of genetical predisposition to atrial fibrillation on CKD or eGFR of the individuals of European ancestry**.

**Supplemental Table 4. Summary statistics of 73 SNPs that were included in the genetic instrument for atrial fibrillation of the individuals of European ancestry in the summary-level MR**.

**Supplemental Table 5. Association between SNPs included in the genetic instrument of European ancestry individuals for atrial fibrillation and the possible confounders in the UK Biobank data**.

**Supplemental Table 6. Summary statistics of 212 SNPs that were included in the genetic instrument for eGFR in the summary-level MR**.

**Supplemental Table 7. Association between SNPs included in the genetic instrument for eGFR and the possible confounders or eGFR (cystatin C) in the UK Biobank data**.

**Supplemental Table 8. Association between SNPs included in the genetic instrument for eGFR in individuals of European ancestry and the possible confounders or eGFR (cystatin C) in the UK Biobank data**.

